# Poliovirus Vaccination Induces a Humoral Immune Response that Cross Reacts with SARS-CoV-2

**DOI:** 10.1101/2021.06.19.21257191

**Authors:** Brittany A. Comunale, Lilly Engineer, Yong Jiang, John C. Andrews, Qianna Liu, Lyuqing Ji, James T. Yurkovich, Roderick A. Comunale, Qiyi Xie

## Abstract

**Background:** Millions have been exposed to SARS-CoV-2, but the severity of resultant infections has varied among adults and children, with adults presenting more serious symptomatic cases. Children may possess an immunity that adults lack, possibly from childhood vaccinations. This retrospective study suggests immunization against the poliovirus may provide an immunity to SARS-CoV-2.

**Methods:** Publicly available data were analyzed for possible correlations between national median ages and epidemiological outbreak patterns across 100 countries. Sera from 204 adults and children, who were immunized with the poliovirus vaccine, were analyzed using an enzyme-linked immunosorbent assay. The effects of polio-immune serum on SARS-CoV-2-induced cytopathology in cell culture were then evaluated.

**Results:** Analyses of median population age demonstrated a positive correlation between median age and SARS-CoV-2 prevalence and death rates. Countries with effective poliovirus immunization protocols and younger populations have fewer and less pathogenic cases of COVID-19. Antibodies to poliovirus and SARS-CoV-2 were found in pediatric sera and in sera from adults recently immunized with polio. Western blot demonstrated antibodies recognized the RNA-dependent-RNA-polymerase (RdRp) of either virus. Sera from polio-immunized individuals inhibited SARS-CoV-2 infection of Vero cell cultures. These results suggest the anti-D3-pol-antibody, induced by poliovirus vaccination, may provide a similar degree of protection from SARS-CoV-2 to adults as to children.

**Conclusions:** Poliovirus vaccination induces an adaptive humoral immune response. Antibodies created by poliovirus vaccination bind the RdRp protein of both poliovirus and SARS-CoV-2, thereby preventing SARS-CoV-2 infection. These findings suggest proteins other than “spike” proteins may be suitable targets for immunity and vaccine development.

## INTRODUCTION

Older adults have an increased likelihood of experiencing more severe COVID-19, compared to children under 18 [1, 2, 3]. Such age-dependent associations were observed during the 2003 SARS-CoV epidemic, as older adults were more likely to contract or die from the disease, compared to younger individuals [4]. This apparent immunity to SARS-CoV and SARS-CoV-2 infection may be associated with childhood vaccinations, such as the poliovirus vaccine. While the poliovirus vaccine has been administered to 90% of the world’s population, antibodies induced by the poliovirus vaccine diminish over time, almost completely by young adulthood [5].

Early clinical studies have shown that some vaccines, including the poliovirus vaccine, can not only protect individuals from the virus for which it was created (polio), but also from other, structurally related viruses [6]. A recent analysis has demonstrated an inverse correlation between susceptibility to SARS-CoV-2 and the severity of COVID-19, and the titers of mumps antibodies [7].

SARS-CoV-2 and the poliovirus are positive ssRNA viruses, whose genomic RNA can be directly used as a protein translation template, as well as for genomic replication driven by an RNA-dependent-RNA-polymerase (RdRp) protein that is translated from the viral template [8, 9]. Due to the importance of the RdRp protein in viral replication, it is the primary target for antivirus drug screening [10]. Thus, the structural similarities in the RdRp of all single-stranded, positive sense RNA viruses may explain the cross-reactivity of polio-immune serum with SARS-CoV-2 antigens [11].

This retrospective study shows that similarities between poliovirus RdRp and SARS-CoV-2 RdRp may explain the apparent protection against SARS-CoV-2 provided by poliovirus vaccination. Two poliovirus vaccine formulations have been used in the worldwide campaign to eradicate polio: an oral live poliovirus vaccine (OPV), and an inactivated poliovirus vaccine (IPV) [12]. OPV is no longer administered in the United States over concerns for the spread of vaccine-derived-poliovirus (VDPV), as there is a remote possibility of VDPV escape into the environment with the live vaccine. IPV, on the other hand, is the form of poliovirus vaccination employed in the United States, due to its zero risk of VDPV, strong efficacy, and minimal risk of side effects. Thus, the study presented here evaluates sera from individuals that were immunized with IPV.

## METHODS

### Relationship between median age and COVID-19 prevalence and mortality

In order to evaluate if there was a relationship between countries’ median age and prevalence or mortality of COVID-19, data on the total number of cases and deaths from the Johns Hopkins University Coronavirus Resource Center were compiled for the 100 countries with the highest prevalence of SARS-CoV-2 [13]. Prevalence and death rates were then calculated for each country based on total population size, as of February 10, 2021. Median age for each country was then included for correlation analyses, calculated using the corrplot package from R Studio software, version 1.4.1103 (https://cran.r-project.org/index.html), between median age and SARS-CoV-2 prevalence and death rates.

### Cloning and expression of SARS-CoV-2 RdRp

The full length *nsp12* gene was cloned into an AmeriDx® *E. coli* over-expression vector pADX75. Similarly, the poliovirus RdRp polyoprotein region 5286bp was amplified to 6564bp (NCBI accession number ALP31139) and cloned into the pADX75 vector. These two vectors were transformed into the BL21 strain respectively. The BL21 strains harboring the vectors were selected and used for protein expression and purification, according to standard protein purification protocols.

### Enzyme-linked immunosorbent assay (ELISA)

An enzyme-linked immunosorbent assay (ELISA) was used for a quantitative analysis of the virus antibody in human sera. ELISA tests were carried out according to standard lab operating procedures. Briefly, the poliovirus vaccine (IPV)’s RdRp full length protein and the SARS-CoV-2 RdRp antigen N protein were coated on the ELISA plate respectively, and the human serum harboring the primary antibody was applied to each well. The primary antibody bound to the antigen coated on the well, and then the horse radish peroxidase (HRP) conjugated secondary antibody, which recognizes the primary antibody, formed a complex with the antigen, antigen-antibody-secondary antibody-HRP. TMB substrate was then added to the ELISA plate. Once the HRP enzyme turned the solution color to blue, the reaction was stopped by adding 2M sulfuric acid, which converted the solution color to yellow. An ELISA reader measured and recorded the activity.

### Western Blot

Recombinant RdRp proteins were separated by SDS-PAGE (Sodium Dodecyl Sulfate Polyacrylamide Gel Electrophoresis), and then were transferred to the PVDF membrane. Each human subject’s serum was applied to the membrane, allowing antibodies in the serum to bind to antigens on the membrane. This binding was captured using a secondary antibody that conjugated with HRP. The HRP’s chemiluminescence substrate then displayed a change of color for the specific RdRp protein band.

### RdRp Polymerase Assay

RdRp activity was determined in vitro by a fluorescence method, which was modified from a technique developed by others for Zika virus RdRp [14]. There was a sample vial and a control vial for each sample, where water replaced ribonucleoside-tri-phosphates (rNTPs). The enzymatic reaction consisted of 0.2M Tris-HCl, pH 8.0, 0.125M NaCl, 40mM MgCl_2_, 10mM spermidine-(HCl)_3_, 2.5uM rNTPs, 2.5mM DTT, and 40nM Syto-82 fluorescence dye. The sample serum was diluted to 1,800 times with 1xPBS, and then 2uL were added to both vials. Samples were incubated at 37°C for 30 minutes. The newly formed double stranded RNA then bound to the Syto-82 fluorescence dye, and emitted 589nm light that could be recorded by a GeneScan™fluorescence reader. The fluorescence signal in each vial was quantified using the mean gray value in an ImageJ program (https://imagej.nih.gov/ij/index.html) [15]. The fluorescence intensity difference between these two vials was designed to reflect the RdRp’s relative activity, such that a positive value represented up-regulated enzymatic activity (normal replication, causing an individual to become ill), and a negative value represented down-regulated enzymatic activity (replication inhibited, preventing viral effects).

### Antiviral Assays

A cytopathic effect (CPE)-based antiviral assay was performed in vivo by infecting Vero-E6-cells in the presence or absence of immune sera to evaluate antiviral activity against SARS-CoV-2 (MEX-BC2/2020) [16]. Vero E6 cells were seeded and incubated for 24 hours. Sera were either pre-incubated first with target cells for one hour at 37°C before infection with SARS-CoV-2, or cells were treated with SARS-CoV-2 for three hours at 37°C to allow viral adsorption, and then the serum specimens were added to the target cells without removing the viral inoculum. The cells were challenged with the viral inoculum and re-suspended in DMEM with 2% FBS (DMEM2). The test sera and virus were maintained in the cell culture media for 96 hours. As a surrogate marker for inhibition of viral replication, inhibition of SARS-CoV-2-induced CPE was measured. Cell viability was monitored with the neutral red (NR) uptake assay. The average absorbance at 540nm (A540) from each well on a 96-well plate was calculated using the following formula:

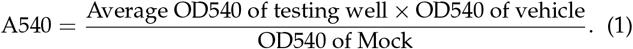

## RESULTS

The objective of this study was to test the hypothesis that immunity against SARS-CoV-2 is dependent upon one’s age, and poliovirus vaccination plays a vital role in this apparent protection (Figure 1). We conducted a population analysis to assess whether there was an association between age and the prevalence and/or mortality of COVID-19 across the globe. Next, we evaluated whether antibodies induced by a childhood vaccine (IPV) may have similarities to SARS-CoV-2 antibodies. Serological tests furthered this exploration through protein quantification, as well as in vitro and in vivo examinations of protein reactivity and inhibition of viral replication of SARS-CoV-2 in polio-immune sera. Data results suggested actionable insights, namely a new possible therapeutic target, RdRp protein, and foundational evidence for a Phase IV clinical trial, which is currently underway.

**Fig. 1.**
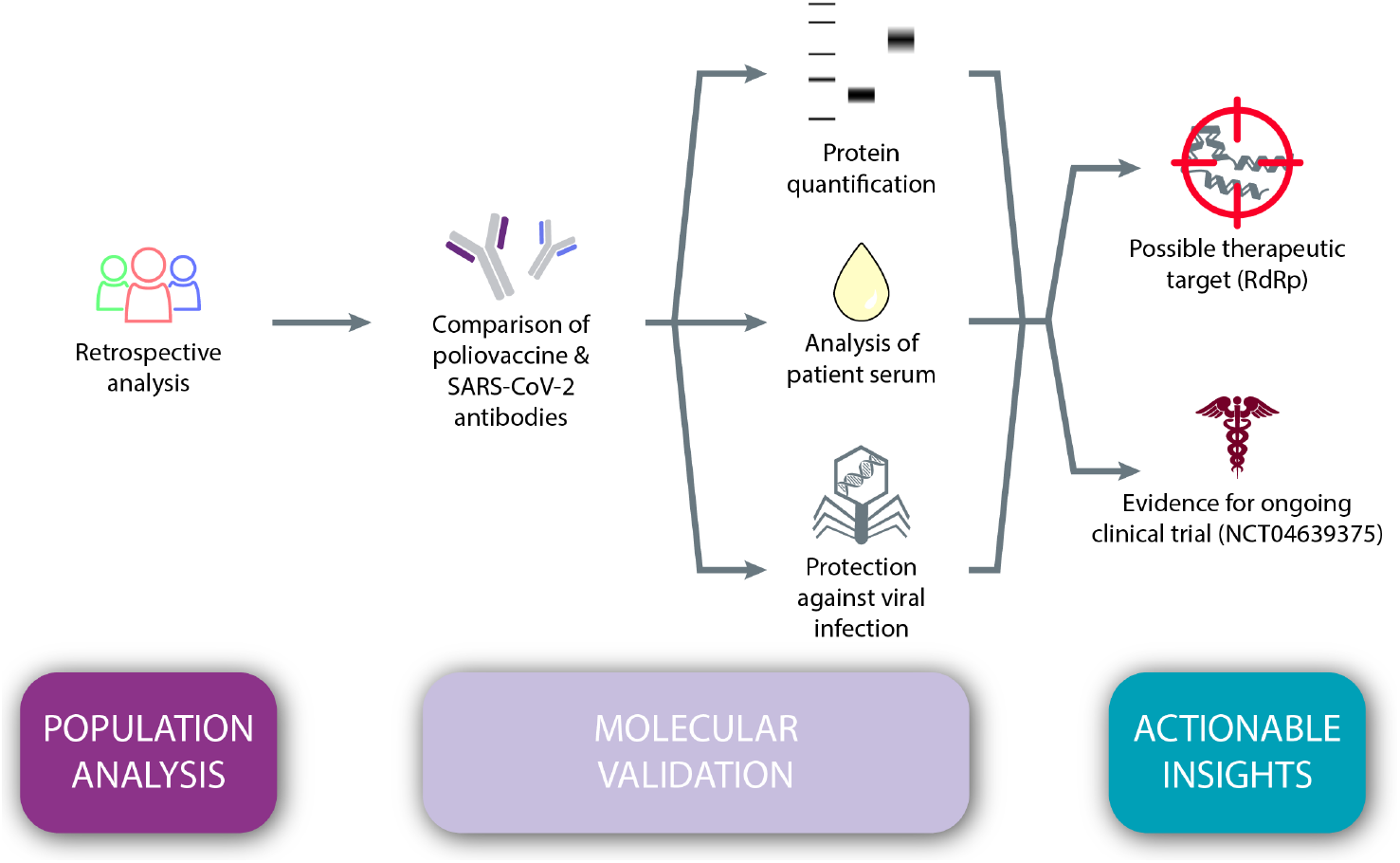
The hypothesis-testing workflow, combining population-level analysis and molecular validation to derive a novel therapeutic target and evidence for an ongoing clinical trial to further validate the hypothesis.

### Relationship between median age and prevalence and/or mortality aspects of COVID-19

We first sought to determine whether there are age-related differences in COVID-19 prevalence and mortality. Using data from the 100 countries with the highest rates of SARS-CoV-2, correlation analyses show there is a positive association between a country’s median age and prevalence rate of SARS-CoV-2 (Pearson *r* = 0.546, *p <* 0.001). There is also a positive correlation between median age and the SARS-CoV-2-related death rate (Pearson *r* = 0.545, *p <* 0.001) (Figure 2). Countries with older national median ages were significantly more likely to have a higher prevalence of SARS-CoV-2, as well as a higher SARS-CoV-2-related death rate.

**Fig. 2.**
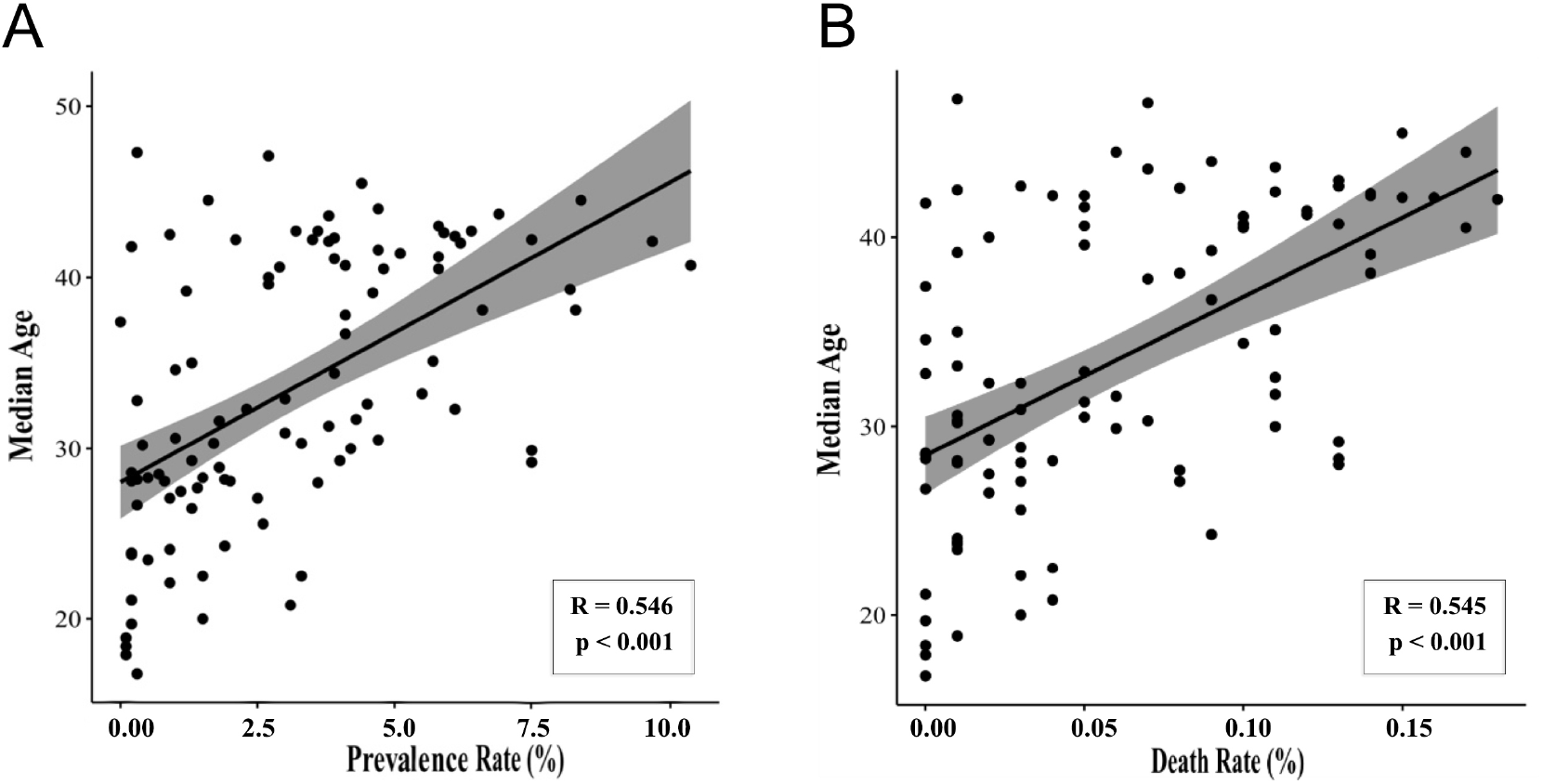
Median age correlates with SARS-CoV-2 prevalence and death rate. Data were collected for the 100 countries with the highest prevalence of SARS-CoV-2 (accessed February 10, 2021). Statistically significant correlations were observed between median age and **(A)** prevalence of SARS-CoV-2 (Pearson *r* = 0.546, *p <* 0.001) and **(B)** COVID-19-related death rate (Pearson *r* = 0.545, *p <* 0.001).

### Poliovirus RdRp shows antibody overlap with SARS-CoV-2 RdRp

We next explored whether antibodies generated by IPV in the human body could recognize SARS-CoV-2 antigens, suggesting COVID-19 infection could be alleviated or prevented by poliovirus vaccination. RdRp was cloned from SARS-CoV-2 and poliovirus, and analyzed by Western blot (Figure 3). The poliovirus RdRp was expressed in bacterial cells and did not produce a full-length version of the protein; the SARS-CoV-2 RdRp was expressed in insect cells and produced the full-length version of the protein. The molecular weight of the full-length RdRp of poliovirus and SARS-CoV-2 were similar, around 130 kD. A truncated form of poliovirus RdRp, produced by bacterial cells, was observed on SDS-PAGE at a molecular weight of around 50-55 kD. Furthermore, the tertiary and quaternary structures of the RdRp were similar and included at least one epitope bound by the RdRp antibody 4E6, as demonstrated by Western blot.

**Fig. 3.**
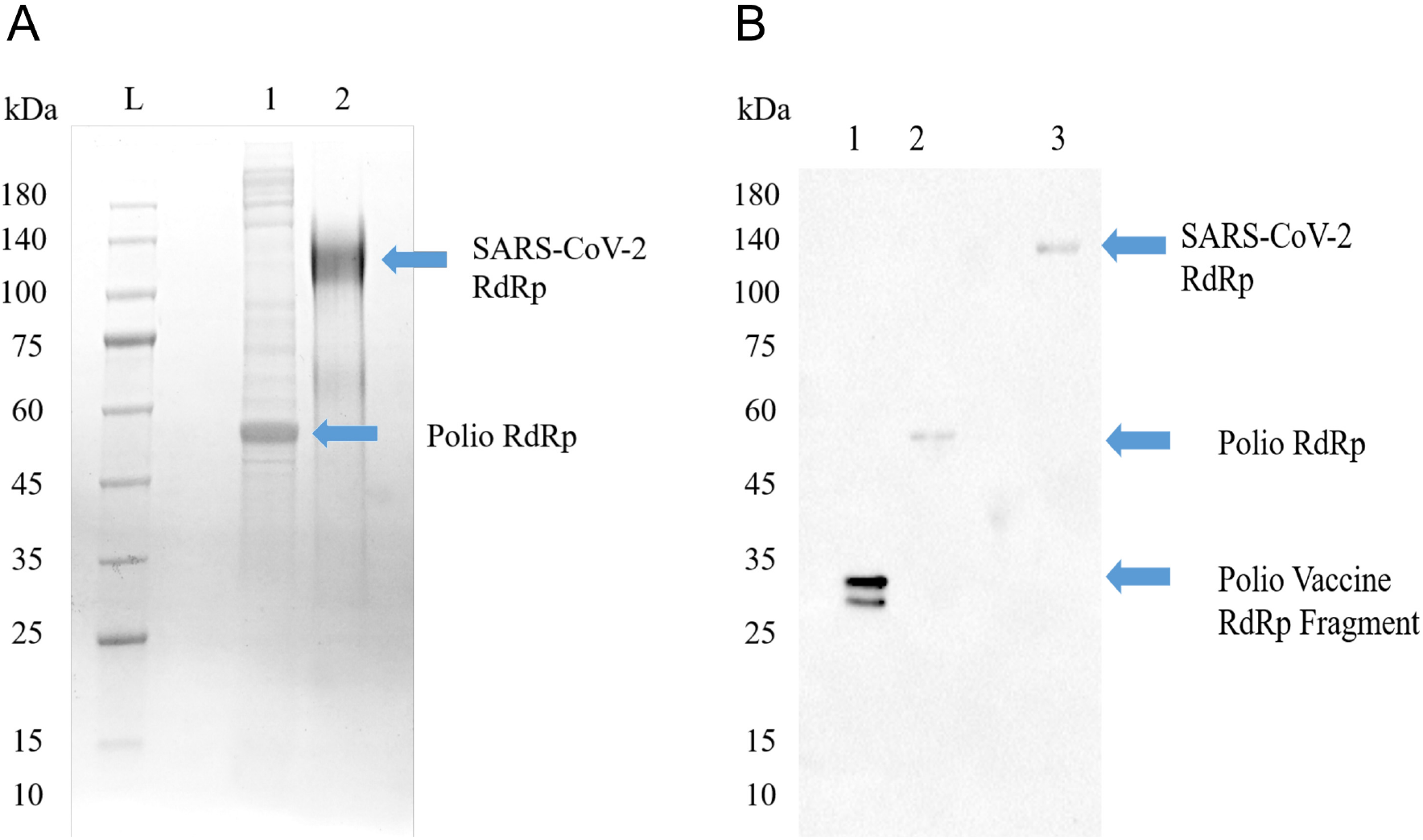
Poliovirus RdRp contains one or more epitopes recognized by SARS-CoV-2 RdRp antibodies. **(A)** SDS-PAGE image of Poliovirus RdRp proteins from bacterial cells (Lane 1) and full SARS-CoV-2 RdRp from insect s2 cells (Lane 2). The poliovirus RdRp from bacterial cells seems to be degraded or not post-translationally modified. Both viral forms of the RdRp in their native configuration have been reported to have a molecular weight around 130 kD. **(B)** Western blot identification of the antigenic activity of RdRp proteins. The primary antibody was a mouse monoclonal antibody to human SARS-CoV-2 RdRp protein, and the secondary antibody was a goat anti-mouse IgG/A/M HRP. Lane 1, a vaccine sample shows a line at about 35 kD; Lane 2, poliovirus RdRp expressed in bacterial cells shows a molecular weight around 50-55 kD, representing an un-modified and possibly truncated form of the RdRp; Lane 3, SARS-CoV-2 RdRp, produced from insect cells, shows a line around 130 kD.

The Western blot shows the poliovirus RdRp. Even though the poliovirus RdRp was expressed in bacterial cells and was not modified post-translation, the cloned poliovirus RdRp contains one or more epitopes recognized by a mouse monoclonal antibody to SARS-CoV-2 RdRp.

### Antibody titers before and after poliovirus vaccination show conferred SARS-CoV-2 immunity

To test the hypothesis that poliovirus vaccination induces an immunity to SARS-CoV-2, we analyzed 204 serum samples from adults (*N* = 123, age 25-89) and children (*N* = 81, age*<* 10) for anti-RdRp antibody titers. Western blot analyses demonstrated antibodies recognized the RdRp of either virus. While the RdRp antigenic activity varied from person to person, sera from those who had been vaccinated for poliovirus showed a significant increase in reactivity to RdRp (*p <* 0.001, Figure 4A). We observed no significant difference in antigenic activity between pediatric and re-immunized adult sera (*p* = 0.85), indicating immunized adults display similar antigenic responses as children that have received their childhood vaccinations (Figure 4B).

**Fig. 4.**
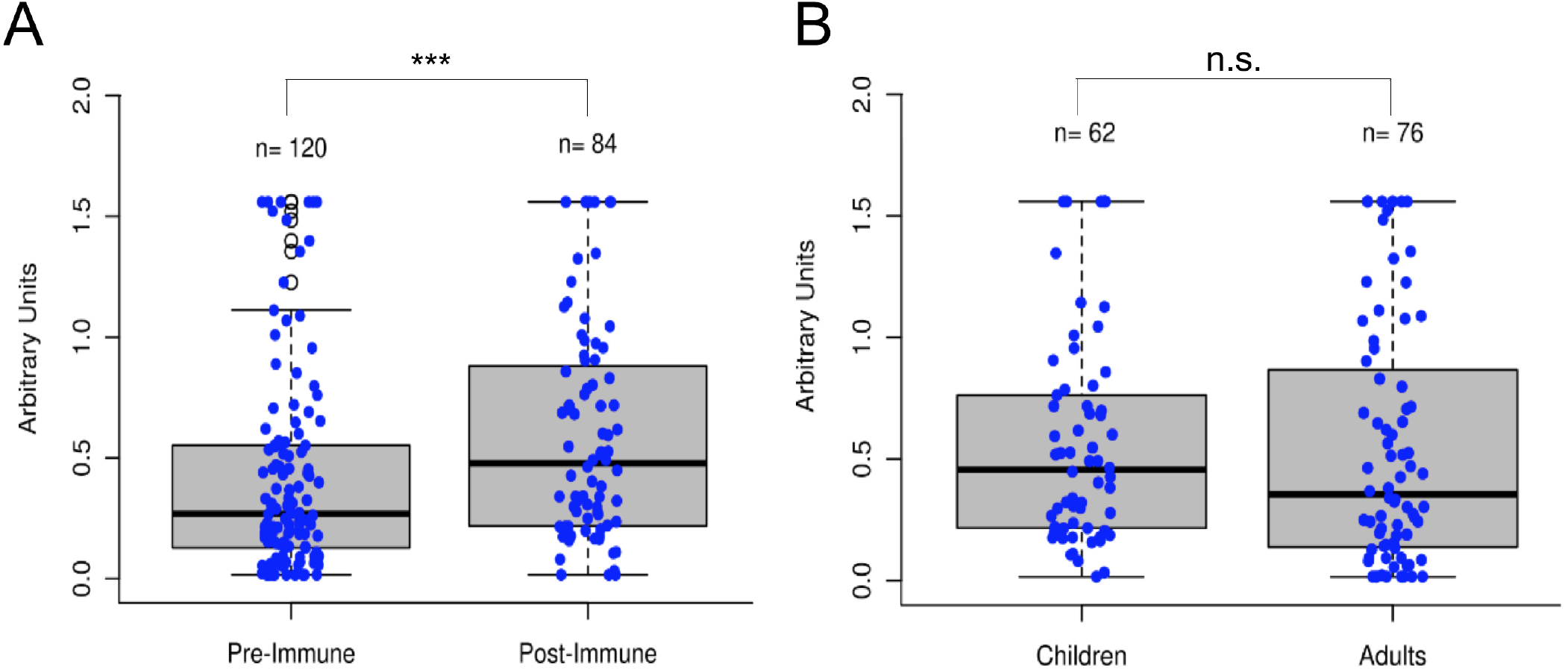
Poliovirus vaccination shows significant increase in RdRp reactivity. **(A)** Boxplot comparison of sera (*N* = 204), before and after poliovirus immunization. Adults significantly gained more antibodies following the poliovirus vaccination, *p <* 0.001. **(B)** Boxplot comparison of pediatric and immunized adult sera. With no significant difference between the two groups (*p* = 0.85), immunized adults display similar antigenic responses as immunized children.

### Antiviral and enzymatic activity of polio-immune sera

Having observed that the poliovirus vaccine significantly affects SARS-CoV-2 RdRp reactivity, our next goal was to understand the mechanism of action behind this phenomenon. Inhibition of SARS-CoV-2-induced cytopathic effect (CPE) in Vero-cell culture was used as a surrogate marker for the inhibition of viral replication. Polio-immune serum demonstrated an antiviral effect in Vero cells at dilutions 1:8 to 1:32, which was stronger when the antisera were pre-incubated with cells, rather than added after viral adsorption. Samples from children fully immunized with IPV (sample #1) and young adults (samples #3A and #3B) show the strongest inhibition of viral CPE when added to cell culture prior to virus (Figure 5). The sample randomly labeled #1 was from pooled sera from children who were on average one years old, collected prior to the onset of the SARS-CoV-2 pandemic; these children would have been immunized against polio as part of national immunization campaigns. The samples randomly labeled as #2 in the figure were pooled from infants who were on average 75 days old; these infants would not have been fully immunized against polio. The remaining samples depicted in Figure 5 were randomly selected from the adult pool of paired samples to demonstrate differences in immunity, based on age and IPV immunization. Sample #3A is from a subject between 25 and 30 years old, who was recently given IPV. Sample #3B is from the same subject after obtaining a second IPV booster. Sample #4A is serum from a subject between 60 and 65 years old pre-IPV vaccination; sample #4B is the post-vaccination serum sample. Sample #5 represents serum from a subject between 13 and 18 years old, who had not recently been vaccinated with an IPV booster.

**Fig. 5.**
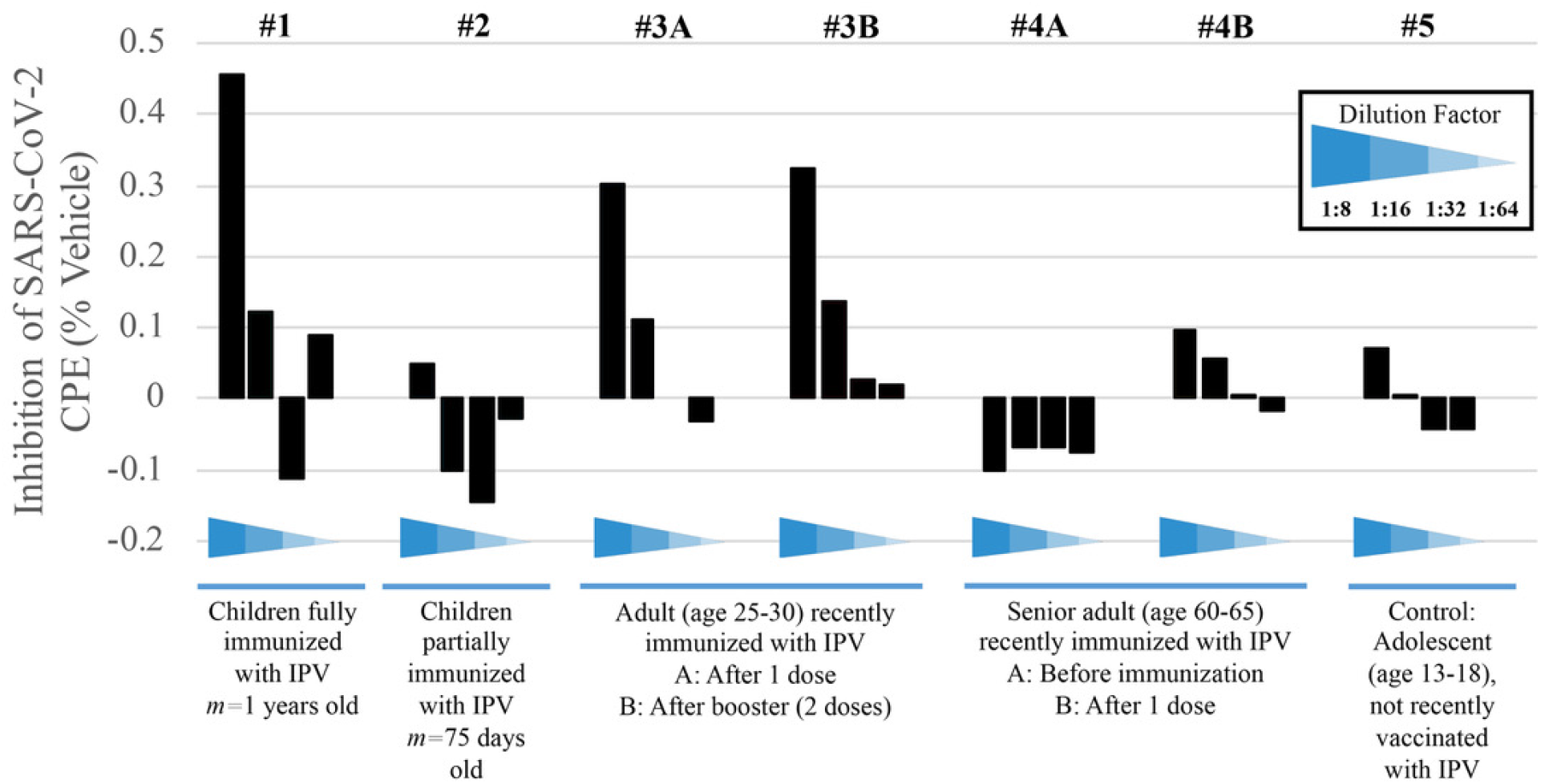
Immunized sera demonstrate higher inhibition of SARS-CoV-2-induced CPE. Sera from five test subjects of varying ages and IPV-immunization stages were randomized and labeled. Sera were added after viral adsorption. Values show the inhibition of the SARS-CoV-2-induced CPE, as a surrogate marker for the inhibition of viral replication.

Children who have received their childhood vaccinations by one year of age display stronger immunity to SARS-CoV-2, compared to children that are less than four months old and still very early in immunization protocols (samples #1 and #2, respectively). This result is expected if, as we propose, poliovirus immunization plays a role in the relative resistance to COVID-19 in younger populations. Samples #3A and #3B further demonstrate how polio immunization (a single inoculation, followed by a second booster two months later, respectively) inhibits SARS-CoV-2-induced CPE.

Protection from poliovirus or SARS-CoV-2 declines as one ages. No inhibition of SARS-CoV-2-induced CPE is evident pre-vaccination (sample #4A), though older adults’ immune systems can respond to an IPV booster (sample #4B). Here, data show approximately a 35% increase of protection from viral CPE, relative to pre-vaccination levels.

While the immunity is lower in the 60-65 year-old adult, compared to the 25-30 year-old adult, IPV provides some protection that the teenager (sample #5), who was not recently vaccinated, does not have.

### Inhibition of RdRp activity is inhibited *in vitro* by polio-immune serum

To explore RdRp as a possible therapeutic target, the effect of polio-immune serum on SARS-CoV-2 RdRp activity was evaluated *in vitro*. Randomly-selected sera from 17 different polio-immunized subjects (7 children; 10 adults) were added to the RdRp reaction mix at a dilution of 1:1800. RdRp activity was then measured by fluorescence signal (Figure 6), with a positive value representing regular enzymatic RdRp activity (virus will replicate normally and cause the individual to become ill) and a negative value representing inhibition of the RdRp enzyme (immunized sera reduce the possibility of viral replication and inhibit the virus from entering the cell). Of the 17 randomly-selected polio-immune sera, 13 (76.5%) were effective in inhibiting the enzymatic activity of RdRp.

**Fig. 6.**
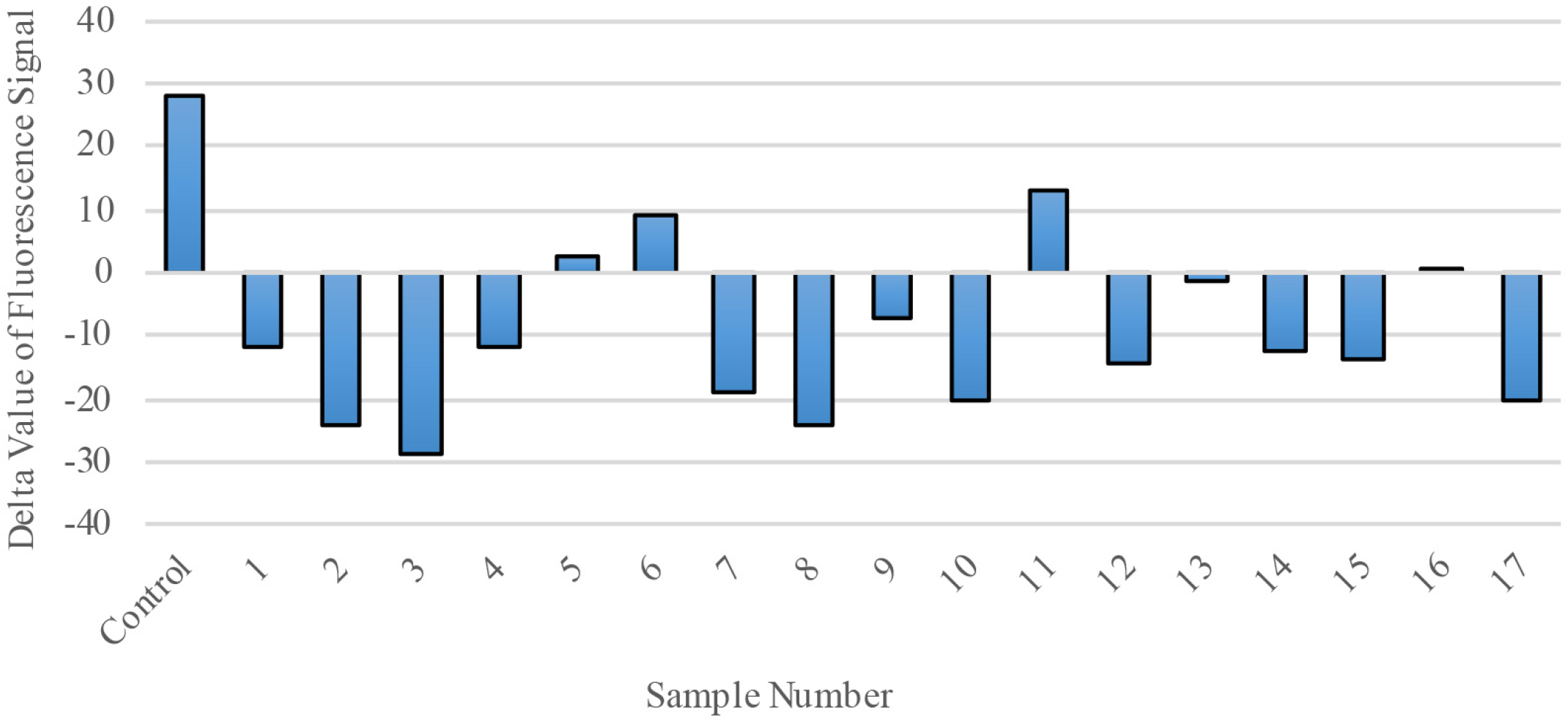
RdRp Enzymatic Activity is blocked by polio-immune serum. RdRp polymerase activity was evaluated by fluorescence staining of *de novo* synthesized RNA. In the presence of diluted serum (1:1800), RdRp’s activity is inhibited. Each bar represents one serum sample (17 randomly-selected samples compared to a control).

## DISCUSSION

With the continued and global spread of SARS-CoV-2, there are open questions regarding the virus’ interaction with the immune system, particularly across various age groups. The interplay between SARS-CoV-2 and other vaccines, such as the poliovirus vaccine, may offer insights into potential therapeutics against COVID-19. Here, we performed a retrospective analysis on sera from 204 individuals to explore the role of poliovirus vaccination in ameliorating the impact of COVID-19 in a population. Our results show poliovirus vaccination raises antibodies that cross-react with SARS-CoV-2, with the primary target of these antibodies being the RdRp of poliovirus and coronavirus. Polio-immune sera, when added to cell cultures before and after viral infection, provide protection from viral CPE. When antisera are added to the RdRp replication system, RNA replication is reduced. Taken together, these results have several important implications.

An IPV-induced adaptive humoral immune response suggests poliovirus immunization in infants and children, as part of national vaccination efforts, provides protection from SARS-CoV-2 infection until young adulthood. The association between national median age and COVID-19 prevalence and mortality rates across countries suggests a lack of immunity to SARS-CoV-2 in older adults, compared to younger individuals, who may still possess immunity from childhood vaccinations, including poliovirus inoculations. Similar demographic patterns were observed from infection prevalence and mortality rates during the 2003 SARS-CoV epidemic [4], where children were less likely to exhibit symptoms related to SARS, compared to older adults [17]. Likewise, such relationships have been evident during the COVID-19 pandemic, as the mortality for people aged 0-24 is substantially less than the mortality in people older than 25 [18, 19, 20]. Childhood vaccinations may have raised antibodies to SARS-CoV-2 in younger individuals, reducing both the prevalence and mortality of COVID-19. Other studies have suggested vaccines that induce an innate immune response, such as BCG, MMR, and poliovirus, may be useful in preventing SARS-CoV-2 [7, 21].

Our results suggest RdRp may be a potential therapeutic target against SARS-CoV-2. We have demonstrated that sera from individuals recently immunized with IPV inhibit SARS-CoV-2-induced CPE in Vero cell culture, and IPV immunization raises antibodies that recognize the RdRp of both poliovirus and SARS-CoV-2. Adults re-immunized with IPV exhibited similar antibody responses to both poliovirus and SARS-CoV-2 RdRp, compared to children who received IPV as part of their childhood vaccinations. Moreover, cell cultures exposed to polio-immune antiserum before live virus exposure were partially resistant to viral entry and showed reduced CPE, compared to pre-immune serum. These findings suggest the anti-RdRp antibodies prevent SARS-CoV-2-induced CPE by inhibiting the cell adsorption or internalization of the virus. This activity seems to depend on the interaction of the viral genome and RdRp, promoting viral entry into the cell. Since prophylactic vaccination is the most effective intervention to protect against infectious diseases, and antibodies raised against SARS-CoV-2 are effective in limiting COVID-19 disease in the majority of patients [22], we suggest IPV immunization may induce adaptive, generally long-term, and specific immunity to poliovirus and SARS-CoV-2 infection. The similarities in structure and function between proteins of SARS-CoV-2 and poliovirus, including RdRp, support this contention.

We propose these results help explain why there is an apparent age-dependent outcome following SARS-CoV-2 infection [1, 2, 3]. There are many exceptions to this observation, as disease is seen in these younger age groups, but at a much lower incidence than the life-threatening disease often seen in adults [23, 24]. While younger individuals may still possess immunity from childhood vaccinations, older adults can mount an adaptive immune response to SARS-CoV-2 following poliovirus re-immunization. Without IPV or OPV boosters, older adults may remain at higher risk of severe disease and mortality [2, 25, 26]. This retrospective study provides a preliminary understanding of the role poliovirus vaccination has in inhibiting viral replication of SARS-CoV-2. We are currently conducting a larger clinical trial (NCT04639375) to provide deeper validation of the potential utility of the safe and effective poliovirus vaccine as a prophylactic measure against COVID-19 infection [27]. The poliovirus vaccine poses minimal risk. The World Health Organization recommends poliovirus vaccine boosters for adults traveling to high-risk zones of polio infection [28]. In comparison to other vaccines that are currently being tested for COVID-19 prevention, poliovirus vaccines, both IPV and OPV, are readily available with prior pharmacological, toxicity, chemical, manufacturing, and control data. Possible risks have been studied for over sixty years and all processes have been well documented and established. Given the current climate of vaccine hesitancy with the COVID-19 vaccine roll-out, and projected vaccine shortage across the globe, the results reported here suggest more attention should be given to a vaccine that 90% of the world’s population received as children. While these data show proteins other than “spike” proteins, such as RdRp, may be suitable targets for immunity and vaccine development, IPV may also be used to complement the new Emergency Use Authorization (EUA) COVID-19 vaccines, to further boost immunity and enhance the population’s health.

This serological study demonstrates poliovirus vaccination produces antibodies that inhibit RdRp function, thereby preventing viral replication that may cause disease progression in infected individuals. Based on these laboratory findings, we conclude polio-vaccinated individuals (children or adults who have recently been immunized with IPV) may have a high level of protection against COVID-19 that non-inoculated individuals do not have.

## Data Availability

Authors can confirm that all relevant data are included in the article.

## ACKNOWLEDGEMENTS

This retrospective study was fully funded by a private source in Southern California. Funders provided finances only and had no involvement in the research activities (project initiation, study design, data collection, analysis, interpretation, results, and inference). All authors had full access to the full data in the study and accept responsibility to submit for publication.

We would like to acknowledge and thank David Cantor and Peter Miller of Cantor BioConnect for providing some serum samples for the retrospective study.

## References

1. Pierce CA, Preston-Hurlburt P, Dai Y, Aschner CB, Cheshenko N, Galen B, et al. Immune responses to SARS-CoV-2 infection in hospitalized pediatric and adult patients. Science Translational Medicine 12 (2020) eabd5487. doi:10.1126/scitranslmed.abd5487.

2. Otto WR, Geoghegan S, Posch LC, Bell LM, Coffin SE, Sammons JS, et al. The epidemiology of severe acute respiratory syndrome coronavirus 2 in a pediatric healthcare network in the united states. Journal of the Pediatric Infectious Diseases Society 9 (2020) 523–529. doi:10.1093/jpids/piaa074.

3. Dong Y, Mo X, Hu Y, Qi X, Jiang F, Jiang Z, et al. Epidemiology of COVID-19 among children in china. Pediatrics 145 (2020) e20200702. doi:10.1542/peds.2020-0702.

4. Leung GM, Hedley AJ, Ho LM, Chau P, Wong IO, Thach TQ, et al. The epidemiology of severe acute respiratory syndrome in the 2003 hong kong epidemic: An analysis of all 1755 patients. Annals of Internal Medicine 141 (2004) 662. doi: 10.7326/0003-4819-141-9-200411020-00006.

5. [Dataset] WHO vaccine-preventable diseases: monitoring system. 2020 global summary (2020).

6. Aaby P, Andersen A, Martins CL, Fisker AB, Rodrigues A, Whittle HC, et al. Does oral polio vaccine have non-specific effects on all-cause mortality? natural experiments within a randomised controlled trial of early measles vaccine. BMJ Open 6 (2016) e013335. doi:10.1136/bmjopen-2016-013335.

7. Gold JE, Baumgartl WH, Okyay RA, Licht WE, Fidel PL, Noverr MC, et al. Analysis of measles-mumps-rubella (MMR) titers of recovered COVID-19 patients. mBio 11 (2020). doi:10.1128/mbio.02628-20.

8. Wolf YI, Kazlauskas D, Iranzo J, Lucía-Sanz A, Kuhn JH, Krupovic M, et al. Origins and evolution of the global RNA virome. mBio 9 (2018). doi:10.1128/mbio.02329-18.

9. Baltimore D. Expression of animal virus genomes. Microbiology and Molecular Biology Reviews 35 (1971) 235–241.

10. Machitani M, Yasukawa M, Nakashima J, Furuichi Y, Masutomi K. RNA-dependent RNA polymerase, RdRP, a promising therapeutic target for cancer and potentially COVID-19. Cancer Science 111 (2020) 3976–3984. doi: 10.1111/cas.14618.

11. Kirchdoerfer RN, Ward AB. Structure of the SARS-CoV nsp12 polymerase bound to nsp7 and nsp8 co-factors. Nature Communications 10 (2019). doi: 10.1038/s41467-019-10280-3.

12. Baicus A. History of polio vaccination. World Journal of Virology 1 (2012) 108. doi:10.5501/wjv.v1.i4.108.

13. [Dataset] COVID-19 dashboard (2021). https://coronavirus.jhu.edu/map.html.

14. Sáez-Álvarez Y, Arias A, del Águila C, Agudo R. Development of a fluorescence-based method for the rapid determination of zika virus polymerase activity and the screening of antiviral drugs. Scientific Reports 9 (2019). doi:10.1038/s41598-019-41998-1.

15. Huang TY, Herr DR, Huang CM, Jiang Y. Amplification of probiotic bacteria in the skin microbiome to combat staphylococcus aureus infection. Microbiology Australia 41 (2020) 61. doi:10.1071/ma20018.

16. Severson WE, Shindo N, Sosa M, Fletcher T, White EL, Ananthan S, et al. Development and validation of a high-throughput screen for inhibitors of SARS CoV and its application in screening of a 100, 000-compound library. Journal of Biomolecular Screening 12 (2006) 33–40. doi:10.1177/1087057106296688.

17. Stockman LJ, Massoudi MS, Helfand R, Erdman D, Siwek AM, Anderson LJ, et al. Severe acute respiratory syndrome in children. Pediatric Infectious Disease Journal 26 (2007) 68–74. doi:10.1097/01.inf.0000247136.28950.41.

18. [Dataset] Roos R. Estimates of SARS death rates revised upward (2003). https://www.cidrap.umn.edu/news-perspective/2003/05/estimates-sars-death-rates-revised-upward.

19. Chan-Yeung M, Xu RH. SARS: epidemiology. Respirology 8 (2003) S9–S14. doi: 10.1046/j.1440-1843.2003.00518.x.

20. Woolf SH, Chapman DA, Lee JH. COVID-19 as the leading cause of death in the united states. JAMA (2020). doi:10.1001/jama.2020.24865.

21. [Dataset] Moyer MW. Could ‘innate immunology’ save us from the coronavirus? (2020). https://www.nytimes.com/2020/05/01/opinion/sunday/coronavirus-vaccine-innate-immunity.html.

22. Zohar T, Alter G. Dissecting antibody-mediated protection against SARS-CoV-2. Nature Reviews Immunology 20 (2020) 392–394. doi:10.1038/s41577-020-0359-5.

23. Viner RM, Whittaker E. Kawasaki-like disease: emerging complication during the COVID-19 pandemic. The Lancet 395 (2020) 1741–1743. doi:10.1016/s0140-6736(20)31129-6.

24. Feldstein LR, Rose EB, Horwitz SM, Collins JP, Newhams MM, Son MBF, et al. Multisystem inflammatory syndrome in u.s. children and adolescents. New England Journal of Medicine 383 (2020) 334–346. doi:10.1056/nejmoa2021680.

25. Grifoni A, Weiskopf D, Ramirez SI, Mateus J, Dan JM, Moderbacher CR, et al. Targets of t cell responses to SARS-CoV-2 coronavirus in humans with COVID-19 disease and unexposed individuals. Cell 181 (2020) 1489–1501.e15. doi: 10.1016/j.cell.2020.05.015.

26. Azkur AK, Akdis M, Azkur D, Sokolowska M, Veen W, Brüggen MC, et al. Immune response to SARS-CoV-2 and mechanisms of immunopathological changes in COVID-19. Allergy 75 (2020) 1564–1581. doi:10.1111/all.14364.

27. [Dataset] E-Mo Biology, Inc. Polio vaccine (IPV) for SARS-CoV-2 and prevention of coronavirus disease (COVID-19) (2020). https://clinicaltrials.gov/ct2/show/NCT04639375.

28. [Dataset] Polio vaccination: For healthcare professionals (2018).

